# Recent changes in COVID-19 Vaccine Hesitancy among Healthcare Workers

**DOI:** 10.1101/2021.03.01.21252457

**Authors:** Emily C. O’Brien, Haolin Xu, Lauren W. Cohen, Elizabeth A. Shenkman, Russell L. Rothman, Christopher B. Forrest, Adrian F. Hernandez

## Abstract

**Introduction:** Early COVID-19 vaccine acceptance rates suggest that up to one-third of HCWs may be vaccine-hesitant. However, it is unclear whether hesitancy among HCWs has improved with time and if there are temporal changes whether these differ by healthcare worker role.

**Methods:** In October 2020, a brief survey was sent to all participants in the Healthcare Worker Exposure Response and Outcomes (HERO) Registry with a yes/no question regarding vaccination under emergency use authorization (EUA): “If an FDA emergency use-approved vaccine to prevent coronavirus/COVID-19 was available right now at no cost, would you agree to be vaccinated?” The poll was repeated in December 2020, with the same question sent to all registry participants. Willingness was defined as a “Yes” response, and hesitancy was defined as a “No” response. Participants were stratified into clinical care roles. Baseline demographics of survey respondents at each timepoint were compared using appropriate univariate statistics (chi-squared and t-tests). Analyses were descriptive, with frequencies and percentages reported for each category.

**Results:** Of 4882 HERO active registry participants during September 1 – October 31, 2020, 2070 (42.4%) completed the October survey, and n=1541 (31.6%) completed the December survey. 70.2% and 67.7% who were in clinical care roles, respectively. In October, 54.2% of HCWs in clinical roles said they would take an EUA-approved vaccine, which increased to 76.2% in December. The largest gain in vaccine willingness was observed among physicians, 64.0% of whom said they would take a vaccine in October, compared with 90.5% in December. Nurses were the least likely to report that they would take a vaccine in both October (46.6%) and December (66.9%). We saw no statistically significant differences in age, race/ethnicity, gender, or medical role between time points. When restricting to the 998 participants who participated at both time points, 69% were vaccine-willing at both time points; 15% were hesitant at both time points, 13% who were hesitant in October were willing in December; and 2.9% who were willing in October were hesitant in December.

**Conclusions:** In a set of cross-sectional surveys of vaccine acceptance among healthcare workers, willingness improved substantially over 2 calendar months during which the US had a presidential election and two vaccine manufacturers released top-line Phase 3 trial results. While improved willingness was observed in all role categories, nurses reported the most vaccine hesitancy at both time points.

---

Vaccine effectiveness at the population level relies on widespread distribution and uptake.^1^ Due to their high risk of exposure to SARS-CoV-2 and essential worker status, the Centers for Disease Control and Prevention (CDC) recommends that healthcare workers (HCWs) receive the first available doses of vaccine.^2^ Early COVID-19 vaccine acceptance rates suggest that up to one-third of HCWs may be vaccine-hesitant.^3,4^ However, it is unclear whether hesitancy among HCWs has improved with time and if there are temporal changes whether these differ by healthcare worker role.

## Methods

The Healthcare Worker Exposure Response and Outcomes (HERO) Registry began in April 2020 to characterize COVID-19 risk factors and outcomes among HCWs in the United States (ClinicalTrials.gov Identifier NCT04342806). Participants self-enroll in an online portal and complete periodic electronic surveys. In October 2020, a brief survey was sent to all registry participants with a yes/no question regarding vaccination under emergency use authorization (EUA):

1. “If an *FDA emergency use-approved* vaccine to prevent coronavirus/COVID-19 was available right now at no cost, would you agree to be vaccinated?”

The poll was repeated in December 2020, with the same question sent to all registry participants. Willingness was defined as a “Yes” response, and hesitancy was defined as a “No” response. Participants were stratified into clinical care roles: registered nurses/licensed practical nurses (RNs/LPNs); physicians; paramedics/emergency medical technicians (EMTs); physician assistants/nurse practitioners (PAs/NPs); and, other health diagnosing/treating practitioners. Analyses were descriptive, with frequencies and percentages reported for each category.

Baseline demographics of survey respondents at each timepoint were compared using appropriate univariate statistics (chi-squared and t-tests). Study oversight was provided by the Duke University Institutional Review Board, and all participants provided informed consent. Data were analyzed with SAS version 9.4.

## Results

Of 4882 HERO active registry participants during September 1 – October 31, 2020, 2070 (42.4%) completed the October survey, and n=1541 (31.6%) completed the December survey. 70.2% and 67.7% who were in clinical care roles, respectively. In October, 54.2% of HCWs in clinical roles said they would take an EUA-approved vaccine, which increased to 76.2% in December (**Figure 1**). The largest gain in vaccine willingness was observed among physicians, 64.0% of whom said they would take a vaccine in October, compared with 90.5% in December. Nurses were the least likely to report that they would take a vaccine in both October (46.6%) and December (66.9%).

**Figure 1.**
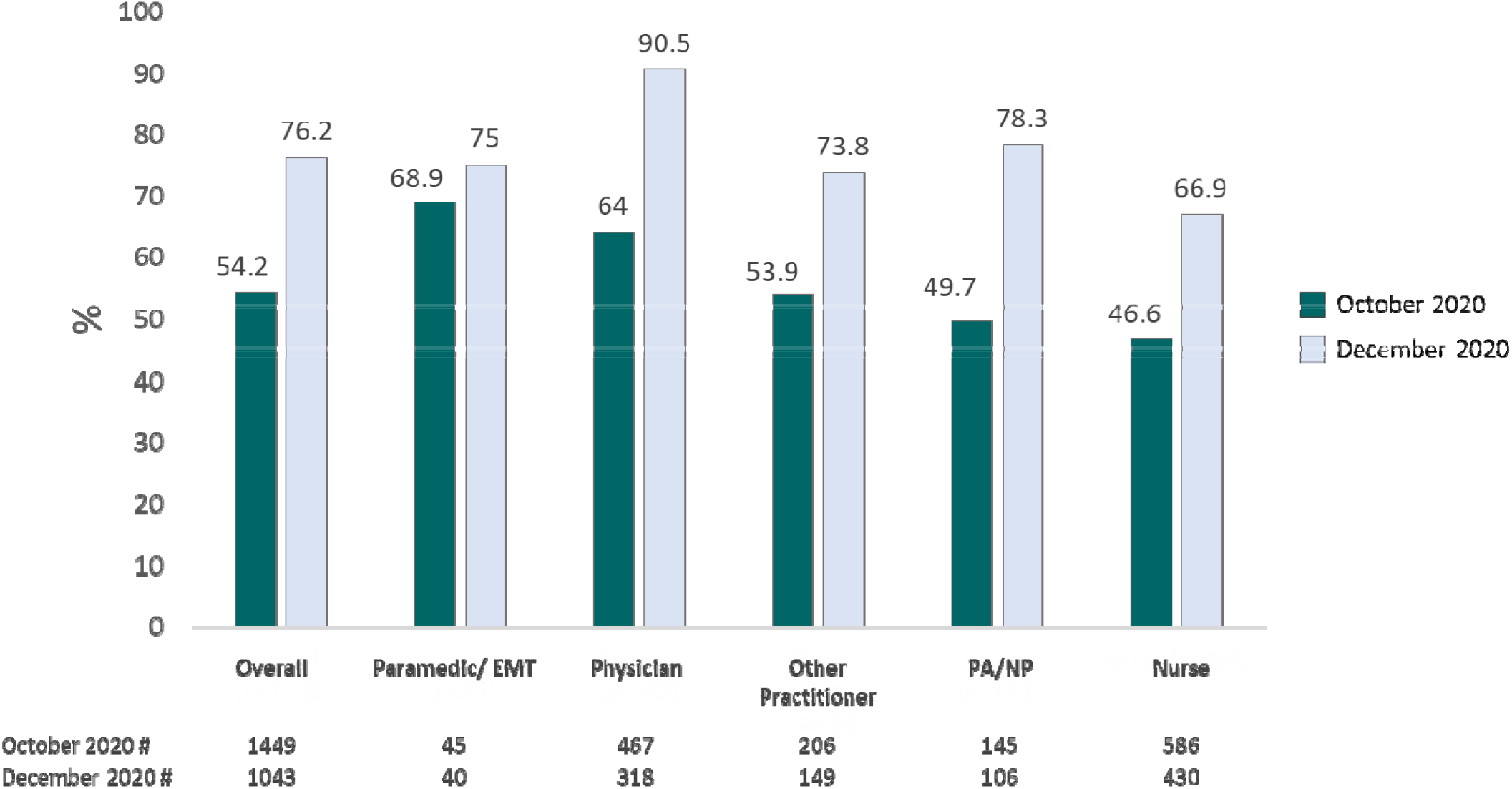
Percent of healthcare workers who would take a COVID-19 vaccine by clinical role in October 2020 and December 2020.

We saw no statistically significant differences in age, race/ethnicity, gender, or medical role between time points. When restricting to the 998 participants who participated at both time points, 69% were vaccine-willing at both time points; 15% were hesitant at both time points, 13% who were hesitant in October were willing in December; and 2.9% who were willing in October were hesitant in December.

## Discussion

In a set of cross-sectional surveys of vaccine acceptance among healthcare workers, willingness improved substantially over 2 calendar months during which the US had a presidential election and two vaccine manufacturers released top-line Phase 3 trial results. While improved willingness was observed in all role categories, nurses reported the most vaccine hesitancy at both time points.

Vaccine hesitancy is a complex problem with context- and time-specific determinants.^5^ Hesitancy exists along a continuum, with corresponding degrees of susceptibility to change.^6^ Our results suggest that COVID-19 vaccine willingness can change substantially with time and with changes in external factors such as experience with actual vaccine administration. Despite temporal improvements, hesitancy remains high among nurses.

Given persistently high national COVID-19 infection rates, our study highlights an urgent need for targeted interventions to increase vaccine acceptance in this critical population especially since healthcare workers are often the most trusted people in the community.

## Data Availability

The data that support the findings will be available in at PCORI-designated repository following study completion.

## Declarations

### Ethics approval and consent to participate

The Duke Institutional Review Board (IRB) provided ethical oversight for the study. All participants provided informed consent prior to data collection.

